# Investigating the role of common *cis*-regulatory variants in modifying penetrance of putatively damaging, inherited variants in severe neurodevelopmental disorders

**DOI:** 10.1101/2023.04.20.23288860

**Authors:** Emilie M. Wigdor, Kaitlin E. Samocha, Ruth Y. Eberhardt, V. Kartik Chundru, Helen V. Firth, Caroline F. Wright, Matthew E. Hurles, Hilary C. Martin

## Abstract

Recent work has revealed an important role for rare, incompletely penetrant inherited coding variants in neurodevelopmental disorders (NDDs). Additionally, we have previously shown that common variants contribute to risk for rare NDDs. Here, we investigate whether common variants exert their effects by modifying gene expression, using multi-*cis*-expression quantitative trait loci (*cis*-eQTL) prediction models. We first performed a transcriptome-wide association study for NDDs using 6,987 probands from the Deciphering Developmental Disorders (DDD) study and 9,720 controls, and found one gene, *RAB2A*, that passed multiple testing correction (p = 6.7×10^−7^). We then investigated whether *cis*-eQTLs modify the penetrance of putatively damaging, rare coding variants inherited by NDD probands from their unaffected parents in a set of 1,700 trios. We found no evidence that unaffected parents transmitting putatively damaging coding variants had higher genetically-predicted expression of the variant-harboring gene than their child. In probands carrying putatively damaging variants in constrained genes, the genetically-predicted expression of these genes in blood was lower than in controls (p = 2.7×10^−3^). However, results for proband-control comparisons were inconsistent across different sets of genes, variant filters and tissues. We find limited evidence that common *cis*-eQTLs modify penetrance of rare coding variants in a large cohort of NDD probands.

## Introduction

Neurodevelopmental disorders (NDDs) such as intellectual disability, epilepsy and autism have a large genetic component^1^. One of the largest studies of NDD patients, the Deciphering Developmental Disorders (DDD) study^2!,3^, consists of 13,451 undiagnosed probands, ∼85% of whom have at least one abnormality of the nervous system, who underwent exome sequencing and exon-resolution microarray analysis. Exome-wide burden analysis has shown that ∼42% of the cases within the cohort are attributable to *de novo* coding mutations in either known or undiscovered Developmental Disorder (DD)-associated genes^4^, with smaller contributions from coding variants following other Mendelian inheritance modes such as X-linked (∼7%)^5^ and autosomal recessive variants (∼3%)^6^. To date, around 41% of probands have received a genetic diagnosis^7^.

Most parents in the DDD study are unaffected; amongst the 1,230 trio probands with an affected father and/or mother, inherited autosomal dominant causes have been identified in 257 (20.9%), which is 2.6% of the 9,859 trio probands^7^. However, there is increasing evidence that incompletely penetrant, inherited, rare, coding variants contribute to risk of NDDs. Firstly, burden analyses have demonstrated that probands with autism have an increased rate of rare deleterious coding variants compared to neurotypical individuals, particularly in a set of ∼3,000 ‘constrained’ genes that are intolerant of loss-of-function (LoF) variation in the general population^8^, and that they over-inherit such variants from unaffected parents^9,10^. Indeed, we find similar signals in our undiagnosed DDD probands and evidence that these variants contribute to risk in a large fraction of probands (Samocha *et al*., manuscript in preparation). Secondly, a small number (N = 22) of DDD probands have been diagnosed with known pathogenic variants in autosomal dominant conditions that were inherited from clinically unaffected parents^11^. In parallel, there is emerging evidence from population-based cohorts that rare, deleterious coding variants in known DD-associated genes^12^ or constrained genes^13,14^ are associated with reduced cognitive function and mental health conditions in the general population. Why these variants are incompletely penetrant represents a major gap in our understanding of DDs and these related phenotypes. Stochastic environmental and genetic modifiers of penetrance likely exist. We previously showed that genome-wide common variants contribute to risk of NDDs^15^; we hypopthesize that at least some of these common variants may act by modifying penetrance of rare coding variants in these disorders.

Castel *et al*. previously presented evidence that *cis*-expression quantitative trait loci (*cis*-eQTLs) modify penetrance of rare coding variants in healthy and disease cohorts^16^. Specifically, they found evidence in a healthy cohort (N = 620) for a depletion of haplotype configurations that should increase penetrance of pathogenic variants (implying a role for negative selection), but that cancer patients (N = 615) and autistic individuals (N = 2,600) were enriched for penetrance-increasing haplotype configurations of pathogenic variants in disease-linked genes. Michaud *et al*. found an example of a similar mechanism in albinism, whereby a common regulatory variant modified the penetrance of two common coding variants in *TYR*^17^. We set out to test whether this mechanism is contributing to the incomplete penetrance of rare, inherited coding variants in DD-associated and constrained genes in the DDD study.

We build on the work of Castel *et al*.^16^ in four main ways. Firstly, we apply more stringent filtering of rare coding variants to focus on those most likely to be damaging. Secondly, we use a cross-tissue, multiple *cis*-eQTL method (UTMOST^18^) to predict gene expression, rather than using a single *cis-*eQTL per gene. Thirdly, we consider genetically-predicted expression in a disease-relevant tissue (cortex) as well as in whole blood, rather than taking the most significant *cis*-eQTL in any tissue for each gene. Finally, we use a within-family design which allows us to avoid potential false positive associations due to population stratification, comparing predicted expression between probands with an inherited rare coding variant to their variant-transmitting parents. Our analysis finds limited evidence to support the hypothesis that *cis*-eQTLs are modifying the penetrance of inherited, putatively damaging coding variants in DDs.

## Results

### Datasets

Individuals from the DDD study were exome-sequenced and genotyped on three different SNP arrays, with some individuals genotyped on more than one array (Supplementary Figure 1). In this work, we used two different array datasets from DDD (see Methods). Analyses on individual NDD probands were based on the dataset used in Niemi *et al*.^15^, comprising 6,987 unrelated NDD cases from DDD with ancestry similar to the 1,000 Genomes^19,20^ Great British samples (henceforth referred to as ‘GBR ancestries’) and 9,270 ancestry-matched controls from the UK Household Longitudinal Study (UKHLS)^21^. These had been genotyped on the Illumina CoreExome chip and imputed to the Haplotype Reference Consortium (HRC) panel^22^. Analyses based on trios used a dataset of 1,700 undiagnosed NDD probands with unaffected parents (of which 1,352 probands were also in the aforementioned CoreExome dataset), all with GBR ancestries, genotyped on either the Illumina OmniExpress chip or the Illumina Global Screening Array and imputed to TOPMed^23–25^.

### Predicting genetically-determined gene expression

To predict the genetically-determined component of gene expression, we used UTMOST^18^, a cross-tissue multi-eQTL method that jointly models multiple tissues when estimating the SNP weights. This has been shown to increase imputation accuracy, particularly for tissues with small sample sizes in the training data, and to generate effective imputation models for an average of 120% more genes than single-tissue methods^18^. We used UTMOST^18^ weights generated from GTEx v6p training data^26^ for two tissues: cortex and whole blood. We chose cortex because it is implicated in various cognitive functions relevant to global developmental delay and intellectual disability^27,28^. We also used weights based on GTEx v6p whole blood (N = 338 versus N = 96 for cortex) in an attempt to balance statistical power with likely physiological relevance to NDDs. While brain tissues may be the most relevant to NDDs, work by Qi *et al*. has shown a gain of power in gene discovery for brain-related phenotypes using blood *cis*-eQTL data on larger sample sizes^29^. We restricted our analyses to genes with cross-validation adjusted p-value < 0.05: 11,103 genes for whole blood, and 11,338 in cortex, with an overlap of 9,476 genes.

### Testing the effect of genetically-predicted gene expression on NDD risk

We first tested whether genetically-predicted expression of any given gene was associated with being an NDD case, regardless of the presence of rare variants, to assess whether *cis*-eQTLs play a role in risk of NDDs when considering average predicted expression. We conducted a transcriptome-wide association study (TWAS) comparing 6,987 unrelated NDD cases with GBR ancestries with 9,270 ancestry-matched UKHLS controls. TWAS have been widely used to try to prioritize likely causal genes underlying complex disease risk^30^. While the GWAS for NDDs in DDD did not identify any genome-wide significant SNPs^15^, a TWAS is generally better powered than a GWAS^31,32^. In our TWAS for NDDs using predicted gene expression from whole blood, we identified one gene passing Bonferroni correction (p = 6.7×10^−7^), *RAB2A* (Figure 1). This gene encodes a protein belonging to the Rab family which is required for protein transport from the endoplasmic reticulum (ER) to the Golgi complex^33^. Mutations in multiple other genes in the Rab family are known to cause NDDs^34–37^. *RAB2A* was not significant in the TWAS in cortex (p = 0.34), and no other genes pass Bonferroni correction (Figure 1). Thus, while *RAB2A* is an interesting candidate for involvement in NDDs, it requires replication in another cohort, and would be more compelling if there were also evidence for association with genetically-predicted expression in a brain tissue or if coding variants in *RAB2A* were associated with in NDDs. Summary statistics for both TWAS can be found in the Supplementary Data.

**Figure 1.**
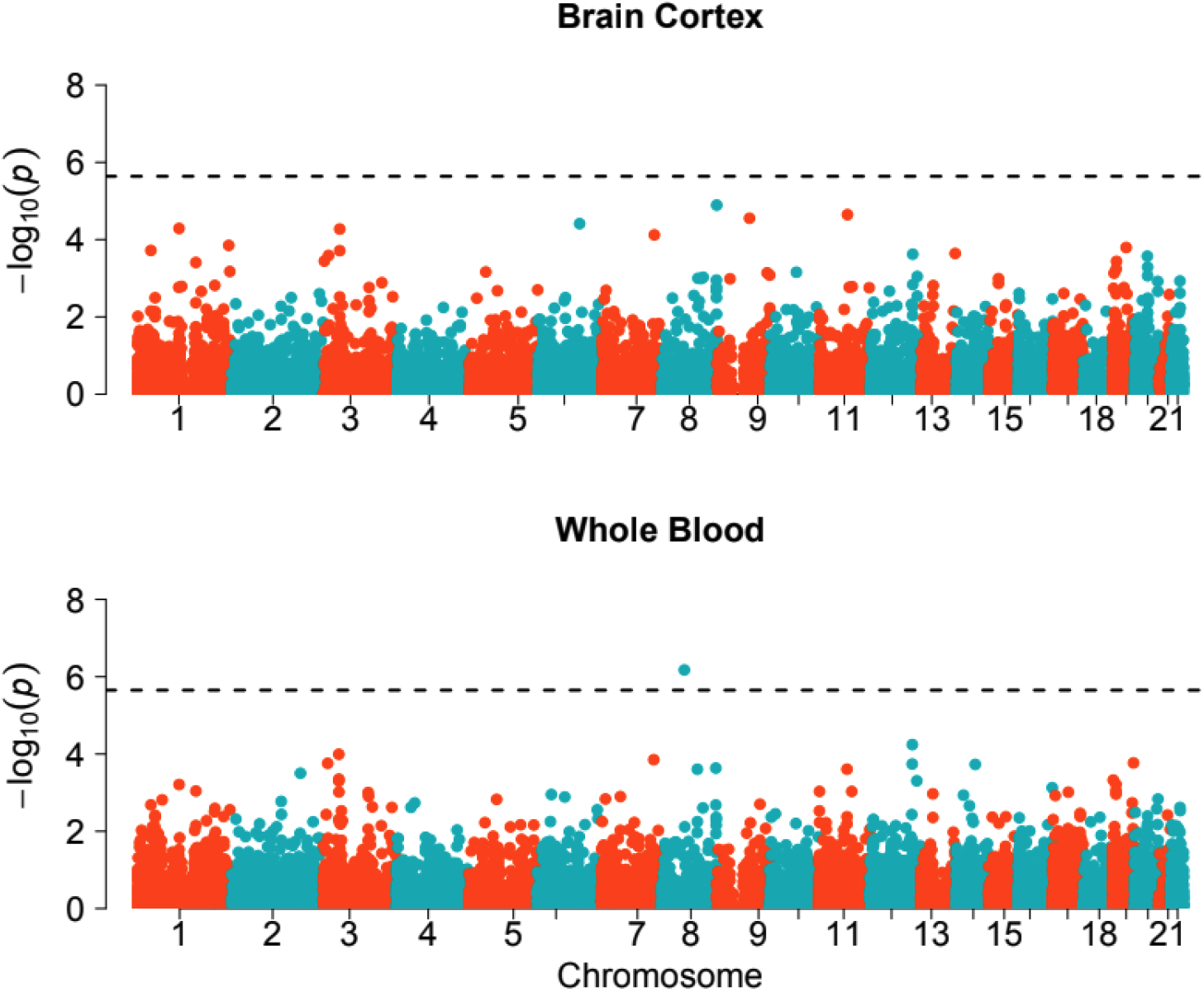
Gene-based p-values from a TWAS comparing genetically-predicted gene expression between 6,987 NDD cases and 9,270 UKHLS controls. The *cis*-eQTL weights are from GTEx v6p cortex (N = 11,338 genes) and whole blood (N = 11,103 genes). Black dotted line represents the significance threshold after Bonferroni correction (p = 0.05/(11,338+11,103)).

The TWAS was intended to assess whether *cis*-eQTLs play a role in risk of NDDs when considering average predicted expression for cases versus controls. We next hypothesized that a small subset of NDD probands might be explained by having an unusual configuration of *cis*-eQTLs for a gene such that it was expressed at an extremely low level (for a LoF mechanism gene) or an extremely high level (for a gain-of-function mechanism gene). Thus, we tested DD-associated genes in the Development Disorder Genotype-Phenotype Database database (DDG2P)^38^ to see whether undiagnosed NDD probands were enriched for extreme genetically-predicted gene expression compared to controls (±3 standard deviations from the mean for controls), using a Fisher’s exact test. None of the genes passed Bonferroni correction (p > 0.05/1,321 DD-associated protein-coding genes = 3.8×10^−5^ in cortex; p > 0.05/1,202 = 4.2×10^−5^ in whole blood).

### Testing whether *cis*-QTLs modify penetrance of rare coding variants in NDDs

We next used the genetically-predicted expression values from NDD probands to test whether *cis*-eQTLs modify the penetrance of putatively damaging, heterozygous, rare coding variants that had been inherited by these probands from their unaffected parents (hereafter: ‘putatively damaging variants’). We focused on rare, inherited heterozygous variants (single nucleotide variants (SNVs) and insertions and deletions (indels); minor allele frequency (MAF) < 1.0×10^−5^ in gnomAD^8^, and ≤ 1.0 × 10^−4^ in DDD) predicted to be damaging, in three categories: i) protein-truncating variants (PTVs) and missense variants (missense badness, PolyPhen-2, and constraint (MPC) ≥ 2^39^) in constrained genes (probability of LoF intolerance (pLI) > 0.9)^40^, ii) PTVs or missense variants in dominant DDG2P genes with a LoF mechanism, and iii) PTVs or missense variants in recessive DDG2P genes with a LoF mechanism. We focused on constrained genes and DD-associated genes with a LoF mechanism because the effect of PTVs and missense variants in such genes is more interpretable (i.e. we assume they result in LoF), whereas identifying which missense variants have an activating or gain-of-function effect in genes for which this is the pathogenic mechanism is more difficult. For the first two categories (constrained genes and dominant DD-associated genes), we hypothesized that the penetrance of the putatively damaging variant is increased by low expression of the other wild-type haplotype (Figure 2A). For the third category (recessive DD-associated genes), we hypothesized that lower expression of the non-variant-carrying haplotype constitutes a ‘second hit’ to the gene, such that, combined with the putatively damaging variant on the other haplotype, gene activity is reduced to a level that is below the pathogenic threshold (Figure 2A).

**Figure 2.**
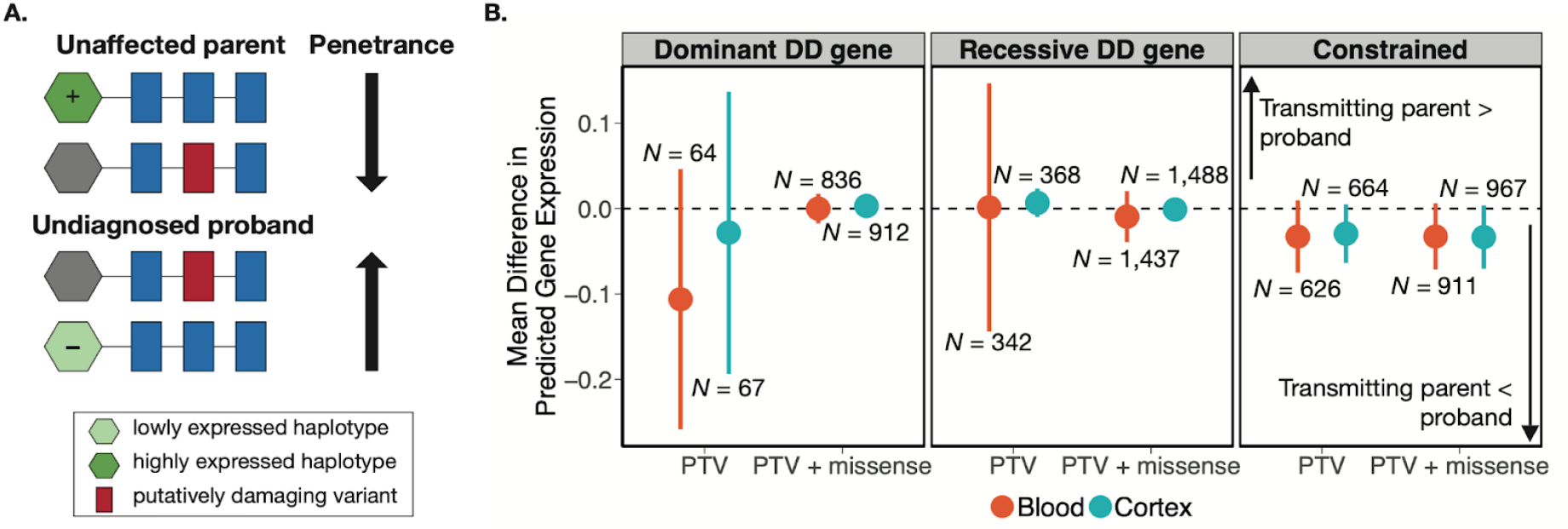
Comparison of predicted gene expression between unaffected variant-transmitting parents and their undiagnosed children with an NDD. A) Schematic figure depicting how *cis*-eQTLs may modify the penetrance of a putatively damaging variant transmitted from an unaffected parent to their undiagnosed child in a gene with a LoF mechanism. The haplotype with the ‘+’ symbol has higher predicted expression based on its *cis*-eQTLs, whereas the one with the ‘-’ symbol has lower predicted expression. B) Mean difference (parent - child) in predicted gene expression between parents transmitting putatively damaging variants and their children with an undiagnosed NDD, with lines indicated 95% confidence intervals. *N* denotes the number of unique child-parent pairs. Predicted gene expression can be interpreted as the inverse quantile-normalised number of reads per kilobase of transcript per million mapped reads (RPKM). The three panels show results for putatively damaging variants in three different sets of genes: dominant DD-associated genes with a LoF mechanism (left), recessive DD-associated genes with a LoF mechanism (middle) or constrained genes (pLI > 0.9) (right). Red and blue dots represent results from genetically-predicted gene expression imputed from whole blood and cortex, respectively. We show estimates considering only PTVs, as well as PTVs and missense variants (with MPC ≥ 2) together.

We began with a within-family test on 1,700 undiagnosed NDD trios to assess whether unaffected parents transmitting a putatively damaging variant were protected by higher genetically-predicted expression of the gene compared to their affected child. Specifically, we ran a one-sided paired *t*-test to compare genetically-predicted gene expression in cortex and whole blood between unaffected parents transmitting a putatively damaging variant and their affected children, with the hypothesis that transmitting parents had higher genetically-predicted expression of the relevant gene. This within-family test controls for population stratification, and allows for a direct comparison of predicted expression of the wild-type haplotype while controlling for both the same putatively damaging variant, and the haplotype wherein it lies (the shared haplotype). We saw no significant difference in genetically-predicted expression between putatively damaging variant-transmitting parents and their children (Figure 2B). We repeated this analysis with a more lenient MAF threshold (MAF < 0.1%) and also saw no significant difference in genetically-predicted expression (Supplementary Figure 2).

We next compared genetically-predicted expression between NDD probands carrying a putatively damaging variant in a given gene with the predicted expression for the same gene in 9,720 UKHLS controls. Specifically, we calculated the percentile ranks of genetically-predicted gene expression values, per gene, in both cortex and whole blood, across undiagnosed NDD probands with putatively damaging variants in the gene and UKHLS controls. We then aggregated these percentile ranks across genes and ran a one-sided Wilcoxon rank test to compare the average ranking of variant-carrying probands with controls. We hypothesized that the probands’ ranked predicted expression values would be lower than in controls. We found that genetically-predicted expression from whole blood of constrained genes harboring putatively damaging variants is lower in variant-carrying probands than in controls (PTVs: p = 1.0×10^−4^; PTVs + missense: p = 2.7×10^−3^, which pass Bonferroni correction for 12 tests) (top right panel of Figure 3; Table 1). We also found nominally significant evidence to suggest that, in cortex, genetically-predicted expression of recessive (p = 0.03) DD-associated genes harboring putatively damaging PTVs is lower in variant-carrying probands than controls (bottom middle panel of Figure 3; Table 1). Similarly, we found nominally significant evidence (p = 0.04) to suggest that, in whole blood, genetically predicted expression of dominant DD-associated genes harboring putatively damaging PTVs and/or missense variants is lower in variant-carrying probands than controls (top left panel of Figure 3; Table 1). These findings are consistent with our hypothesis that the haplotype with the wild-type allele may be expressed at a lower level, thus increasing the penetrance of putatively damaging variants in undiagnosed NDD cases compared to controls. However, results were inconsistent across the two tissues and gene sets tested. Furthermore, this analysis does not take into account whether any controls carry a rare, potentially damaging variant in the same gene as the cases (since sequence data are not available for controls), and is thus less robust than the within-family analysis mentioned above. We repeated this analysis with a more lenient MAF threshold (MAF < 0.1%) and found similarly inconsistent results (Supplementary Figure 3).

**Table 1.** Results of one-sided Wilcoxon rank test for percentile-ranked predicted expression of genes harboring putatively damaging variants in undiagnosed NDD probands compared to controls. The sample size for each test is the (number of probands with a putatively damaging variant) + (number of unique genes in which a proband has a putatively damaging variant x N controls (9,270)).

| Tissue | Variant Type | Gene Type | N genes | N cases with variant | Mean case rank | Mean control rank | P-value |
| --- | --- | --- | --- | --- | --- | --- | --- |
| Blood | PTV | dominant DD | 43 | 85 | 57.82 | 50.00 | 0.99 |
| Blood | PTV + missense | dominant DD | 156 | 1,055 | 48.68 | 50.00 | 0.04 |
| Cortex | PTV | dominant DD | 50 | 90 | 45.85 | 50.00 | 0.08 |
| Cortex | PTV + missense | dominant DD | 186 | 1,168 | 49.54 | 50.00 | 0.26 |
| Blood | PTV | recessive DD | 297 | 469 | 48.40 | 50.00 | 0.10 |
| Blood | PTV + missense | recessive DD | 590 | 1,895 | 50.38 | 50.00 | 0.80 |
| Cortex | PTV | recessive DD | 316 | 501 | 47.72 | 50.00 | 0.03 |
| Cortex | PTV + missense | recessive DD | 627 | 1,950 | 50.11 | 50.00 | 0.61 |
| Blood | PTV | constrained | 386 | 901 | 46.79 | 50.00 | $1.0 \times 10^{-4}$ |
| Blood | PTV + missense | constrained | 731 | 1,242 | 48.11 | 50.00 | $2.7 \times 10^{-3}$ |
| Cortex | PTV | constrained | 440 | 951 | 49.61 | 50.00 | 0.32 |
| Cortex | PTV + missense | constrained | 839 | 1,327 | 49.85 | 50.00 | 0.41 |

**Figure 3.**
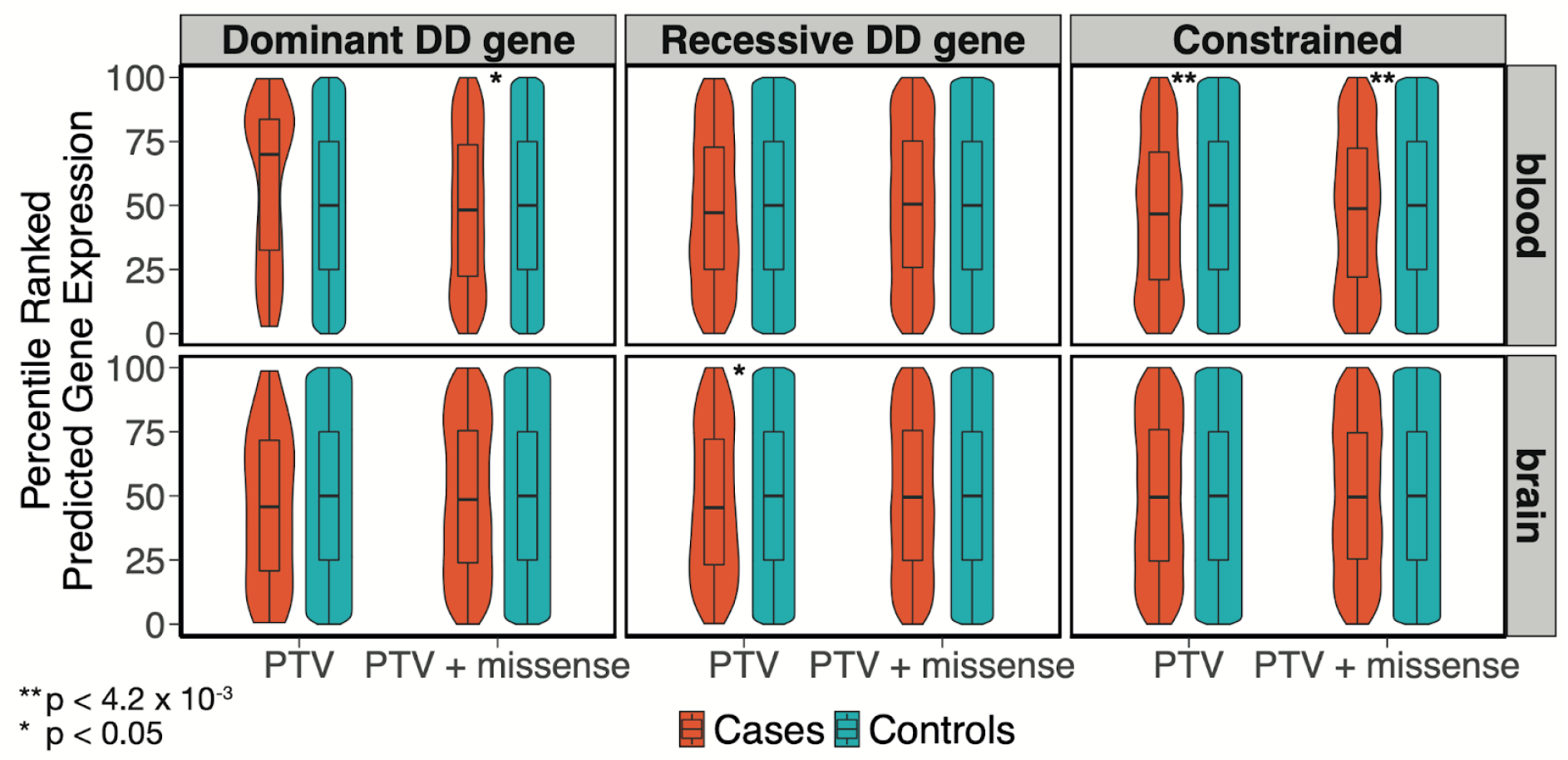
Violin and box plots of percentile-ranked genetically-predicted expression values of genes harboring putatively damaging variants in undiagnosed NDD cases, compared to controls. Vertical lines of the box plot indicate the range and horizontal lines indicate the lower quartile, median and upper quartile. The p-value is from a one-sided Wilcoxon test assessing whether cases are lower than controls. The Bonferroni multiple testing threshold is p = 0.05/12 = 4.2×10^−3^.

A limitation of these analyses is that, while these variants were filtered to be rare and predicted to be damaging by *in silico* predictors, many of the variants are likely not damaging, or only have mild effects. Thus, we investigated differences in predicted gene expression for specific cases in which the proband had a diagnostic variant that was inherited from an unaffected parent, and thus incompletely penetrant. We focused on a set of twenty-two variants in DDG2P genes that were known to be pathogenic based on their ClinVar annotation and that were deemed pathogenic/likely pathogenic by the proband’s clinician, despite having been inherited from an unaffected parent^11^. We postulated that this set of variants is the most likely to show evidence of this mode of modified penetrance. In Table 2, we show the results for the five variants that fell in genes with a predicted loss-of-function consequence and whose expression was predicted by UTMOST with FDR-adjusted p-value < 0.05 in blood and/or cortex. For three of these variants, our hypothesis was supported by the results based on the one tissue for which predicted expression was available (*RORA*, plus two variants in *EBF3*). However, there were two variants (those in *ANKRD11* and *NF1*) for which results were inconsistent between genetically-predicted gene expression values from cortex versus from whole blood. This is either because one or more *cis-*eQTLs have different predicted directions of effect in the two tissues, or different *cis-*eQTLs are used to predict expression in the two tissues, or some combination of the two. Thus, even in this small set of variants for which we most expected to see some signal of this mode of modified penetrance, the evidence for it is inconsistent.

**Table 2.** Comparison of genetically-predicted gene expression in cortex and whole blood between variant-transmitting parents and their children, for a set of known pathogenic variants from ClinVar that were deemed pathogenic/likely pathogenic in the proband by their clinician, despite being inherited from an unaffected parent^11^. These genes are listed in DECIPHER^41^ as causing DDs via a LoF^41^. In the two rightmost columns, NA indicates that the gene’s expression was not sufficiently well predicted in that tissue to be considered.

| Gene | Location (GRCh37)<br>(chr:pos:ref:alt) | Gene<br>consequence | Variant consequence | Child vs<br>parent<br>(Blood) | Child vs<br>parent<br>(Cortex) |
| --- | --- | --- | --- | --- | --- |
| *EBF3* | chr10:131665510:G:<br>A | loss-of-function | NM_001005463.3:p.Arg303* | child lower | NA |
| *EBF3* | chr10:131665510:G:<br>A | loss-of-function | NM_001005463.3:p.Arg303* | child lower | NA |
| *RORA* | chr15:60789728:G:A | loss-of-function | NM_134260.2:p.Arg533* | child lower | NA |
| *ANKRD11* | chr16:89348863:G:A | loss-of-function | NM_001256182.2:p.Arg1363<br>* | child higher | child lower |
| *NF1* | chr17:29560229:T:C | loss-of-function | NM_001042492.3:p.Trp1236<br>Arg | child higher | child lower |

## Discussion

In this work, we evaluated whether levels of gene expression predicted based on common variants modulated NDD risk and penetrance of rare, inherited damaging variants in a large sample of probands from the DDD study. In a TWAS comparing NDD cases with controls, we found one gene passing multiple testing correction in whole blood, *RAB2A*. We are cautious in interpreting this result for several reasons: there is no additional supporting evidence in the literature, the gene showed no signal in cortex (a more disease-relevant tissue), it has not yet been replicated in an independent sample, and TWAS hits may not reflect the true causal gene^30,42^. In evaluating the role of *cis*-eQTL-mediated gene expression in modifying penetrance of rare, inherited, damaging variants, our within-family test found no evidence of this, while results from a case/control analysis were more equivocal, supporting our hypothesis for some gene set-tissue-variant type combinations but not others. Analysis of a small set of known pathogenic, incompletely penetrant variants also failed to provide consistent evidence that their penetrance was being modified by *cis*-eQTLs.

There are several limitations to our analysis. A major one is that, in an attempt to boost power, we aggregated evidence across rare variants in many genes, many of which are likely not deleterious. We used stricter filtering of rare coding variants than Castel *et al*.^16^, focusing on a set that is over-transmitted from unaffected parents to probands in DDD (Samocha *et al*., manuscript in preparation). For example, we used a more stringent MAF filter of < 0.001% in gnomAD^8^ rather than MAF < 1%. Castel *et al*. considered all missense variants with CADD > 15 and, at least for part of their analysis, assumed that penetrance would be increased by higher expression of the variant-containing haplotype. This is not what one would expect if the variant results in loss-of-function, and hence, we restricted to variants that seemed more likely to have a LoF consequence (PTVs or missense variants in genes constrained against LoF variation and/or with a known LoF disease-causing mechanism), and considered predicted expression of the other haplotype. Despite our more stringent filtering, many of the rare variants we included still likely do not result in true LoF, which undoubtedly reduces our power.

Another limitation is that our set of NDD probands is phenotypically heterogeneous; 88% of recruited DDD probands also had abnormalities in at least one other organ system^15^. This makes it challenging to choose an appropriate tissue in which to predict gene expression, since this may differ between probands. Furthermore, eQTLs can be cell-, state- and time-dependent^30,43–49^, and the more relevant cell type and developmental stage to consider is even more difficult to pinpoint, and likely will differ between probands. It may be that selecting *cis*-eQTLs ascertained in fetal brain would be more physiologically relevant for neurodevelopmental disorders than those from adult brain.

A fundamental problem is that common *cis*-eQTLs only explain about 10% of the genetic variance in real gene expression^30^, which limits their predictive accuracy. For example, using a single tissue method, PrediXcan, the average Pearson correlation between predicted gene expression and real gene expression across tissues is around 0.14^50^. UTMOST^18^ modestly improves average imputation *r*^2^ across tissues over PrediXcan by 36.8%^18^. Moreover, genes associated with Mendelian diseases are likely depleted for common *cis*-eQTLs^40,51–55^. Future studies could potentially incorporate methods that use rare eQTLs^53^, *trans*-eQTLs, and epigenetic information^56^ to predict gene expression. Alternatively, they could measure gene expression directly using RNA sequencing, to assess whether expression patterns (whatever their causes) are modifying penetrance of rare variants. To have sufficient power, such studies would either need to be very large or targeted at individuals with putatively pathogenic transmitted variants.

In conclusion, we did not find strong evidence to support the hypothesis that common *cis*-eQTL-mediated gene expression modifies NDD risk or penetrance of rare coding variants in NDDs. Despite addressing this in one of the largest available datasets of NDD probands, our power was still limited by the phenotypic heterogeneity of the cohort, uncertainty about which variants have true effects, and the low accuracy of gene expression prediction models. Future studies should consider this hypothesis in larger datasets with direct measurements of expression and genome sequencing data to evaluate rare variants that could alter gene expression. They should also consider alternative explanations for this apparent incomplete penetrance of rare inherited variants, such as a modifying role of polygenic background^57^, epistasis, stochastic effects, alternative splicing, changing effects of these rare variants with age, or environmental factors.

## Methods

### Preparation of DDD cases and UKHLS controls on the CoreExome chip

We focused the case-control analyses (Figure 1, Figure 3, Table 1) on the DDD and UKHLS data that were used in Niemi *et al*.^15^. These included 6,987 unrelated NDD cases from DDD with GBR ancestries (defined based on their clustering around the 1000 Genomes Great British samples) and 9,720 ancestry-matched controls from UKHLS. These samples were genotyped on the Illumina HumanCoreExome chip^15^. Pre-imputation quality control of these genotype data and imputation to the HRC panel are described in Niemi *et al*.^15^. Post-imputation, genotype data were filtered to SNPs with an imputation r^2^ ≥ 0.8 and MAF > 1%.

### Preparation of genotype data from DDD trios

The analyses in Figure 2 and Table 2 are based on a set of DDD trios that had been genotyped on either the Illumina Infinium Global Screening Array (GSA) or the Illumina OmniChipExpress chip. The preparation of those data is detailed below, with a summary of the filtering steps in Supplementary Figure 4.

#### Quality control and imputation of the GSA data

9,850 DDD samples were genotyped on the GSA at King’s College London in March 2020. Samples were genotyped in a pilot batch (N = 1,152), and a second, larger batch (N = 8,698). Tables S1 and S2 show the results of the quality control steps applied to samples and SNPs before and after merging the batches, respectively.

Samples were checked for concordance with whole-exome sequencing (WES) data previously generated and cleaned on all DDD individuals, described in previous publications^2^; discordant samples were removed, as were sample swaps and duplicate samples. Individuals with ≥ 5% SNPs missing genotyped data were removed. After examining the heterozygosity rate per individual versus the proportion of missing genotypes per individual^58^, we removed individuals with a heterozygosity rate below 0.158 and above 0.17. Trios for which the offspring had > 200 Mendelian errors (∼ 0.03% error rate) were removed.

Palindromic, duplicated and multiallelic markers were removed, as well as indels. Markers with either a call rate < 5%, a MAF < 1%, or with significant deviation from Hardy-Weinberg Equilibrium (p < 1.0 ×10^−6^) were also removed. Markers with a significantly different non-missing rate (p < 1.0 × 10^−50^) or marked allele frequency difference between the pilot batch and second batch of GSA data were removed. SNPs with Mendelian errors in > 1% of trios were removed. This left 9,534 individuals and 474,926 genotyped SNPs before imputation.

After this SNP-level QC, we identified individuals of GBR ancestries. (See Supplementary Figure 5 and Supplementary Methods for further detail). This left 8,879 individuals.

Imputation was carried out using the TOPMed imputation server. After removing variants with imputation *r*^2^ ≥ 0.8, 35,901,148 autosomal SNPs remained.

#### Preparation of DDD trios genotyped on the Omni chip

Niemi *et al*.^15^ also made use of a set of 3,504 individuals from DDD who had been genotyped on the Illumina OmniChipExpress chip. The pre-imputation quality control of these genotype data has been described previously^15^. The prior study used the Haplotype Reference Consortium (64,976 low-coverage genomes) as an imputation panel. We re-imputed the post-QC genotype data using the TOPMed reference panel (97,256 high-coverage genomes) and imputation server, which uses Eagle2 for phasing and minimac4^24^ for genotype imputation^23–25,59^. We removed SNPs with imputation *r*^2^ < 0.8, leaving 36,904,864 SNPs.

#### Merging and checking ancestry of the DDD trios genotyped on the Omni and GSA chips

We merged the data from the GSA and Omni chips (11,227 individuals) and verified that the individuals were well-matched for ancestry (Supplementary Figure 6). See Supplementary Methods for further details. This merged dataset contained 3,344 trios of which all three individuals were inferred to have GBR ancestries.

#### Subsetting DDD trios for analyses

We then removed trios in which probands or parents were related to individuals in other trios up to three degrees of relatedness. To identify related individuals across trios, we ran the –genome command in PLINK v1.9^60^. Pairs of individuals with 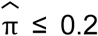 were considered unrelated. After filtering, we retained a set of 3,170 unrelated trios with GBR ancestries. Among these, 2,422 probands were considered undiagnosed (see section below on ‘Identifying undiagnosed probands’), and 2,002 had unaffected parents. Finally, of those, 1,700 had a neurodevelopmental disorder, defined as having one of the following HPO terms^61^: abnormal metabolic brain imaging by MRS (HP:0012705), abnormal brain positron emission tomography (HP:0012657), abnormal synaptic transmission (HP:0012535), abnormal nervous system electrophysiology (HP:0001311), behavioural abnormality (HP:0000708), seizures (HP:0001250), encephalopathy (HP:001298), abnormality of higher mental function (HP:0011446), neurodevelopmental abnormality (HP:0012759), abnormality of the nervous system morphology (HP:0012639). This filtering process is depicted in Supplementary Figure 4.

### Identifying undiagnosed probands

The DDD exome analysis team identified potentially clinically relevant variants from the WES and arrayCGH data as described in Wright *et al*.^3^. The clinical filtering procedure focuses on identifying rare damaging variants in a set of genes known to cause developmental disorders (DDG2P) (https://www.deciphergenomics.org/ddd/ddgenes), that fit an appropriate inheritance mode. Variants that pass clinical filtering are uploaded to DECIPHER^41^, where the probands’ clinicians classify them as either ‘definitely pathogenic’, ‘likely pathogenic’, ‘uncertain’, ‘likely benign’ or ‘benign’. We downloaded all DDD variants from DECIPHER^41^, on July 30, 2021. Of these, 23.5% had not yet been classified by clinicians. Thus, to better differentiate between diagnosed and undiagnosed probands, we estimated positive predictive values (PPV) for different classes of variants and used this to identify probands for whom the variants that passed clinical filtering seemed likely to contain the true diagnosis. We estimated positive predictive values as the proportion of variants in that class (e.g. *de novo* PTV in dominant gene with a loss-of-function mechanism) that clinicians had rated as ‘pathogenic’ or ‘likely pathogenic’. The classes of variants considered and their positive predictive values are shown in Supplementary Table 3.

We defined ‘undiagnosed probands’ as those that did *not* fulfill at least one of the following criteria:

i. the proband was amongst the diagnosed set in a thorough reanalysis of the first 1,133 trios^62^,
ii. the proband had at least one variant (or pair of compound heterozygous variants) rated as pathogenic’ or ‘likely pathogenic’ by a clinician,
iii. the proband had at least one variant (or pair of compound heterozygous variants) in a class with a high or medium PPV (i.e. PPV>50%; see Supplementary Table 3) that passed clinical filtering but had not yet been rated by clinicians,
iv. the proband had a *de novo* PTV in a gene with a pLI > 0.9^40^.

### Predicting gene expression

We used SNP weights from UTMOST^18^ to genetically predict gene expression based on the imputed genotype dosage files. UTMOST^18^ is a cross-tissue gene expression imputation model^18^. Genetically-predicted expression was only generated for genes which had a cross-validation FDR-adjusted p-value < 0.05 in the dataset used to build the models. We used *cis*-eQTL SNP weights generated from two datasets: GTEx v6p brain cortex and whole blood.

### Transcriptome-wide association study (TWAS) for NDDs

We ran two TWASs using predicted gene expression with weights derived from the GTEx v6p brain cortex (N = 96) and GTEx v6p whole blood (N = 338). We predicted gene expression using estimated SNP weights from UTMOST, then ran logistic regression of predicted expression for each gene on case status (N = 6,987 cases, N = 9,270 controls), controlling for the first 10 genotype PCs. We set a Bonferroni significance threshold of p-value < 2.23 × 10^−6^ for the two TWAS (0.05/(11,338 genes in cortex + 11,103 genes in whole blood)).

### Quality control of whole-exome sequencing data

A brief overview of the quality control carried out on the DDD whole-exome sequencing data can be found in Supplementary Table 5. We focused on SNVs and indels. Coding consequences are defined by the worst annotation across transcripts using the Variant Effect Predictor^63^.

When multiple indels are found nearby in the same individual, this frequently indicates a complex mutational event. Properly resolving these complex mutational events would require haplotype-aware annotation, which was beyond the scope of this work. Consequently, we removed instances in which a sample had more than one indel in a given gene. This filter removed fewer than 4% of all indels with a MAF < 1% in our dataset.

### Investigating role of genetically-predicted gene expression in modifying penetrance of rare variants

Amongst the undiagnosed, NDD probands of GBR ancestries with unaffected parents, we identified those with at least one rare (MAF < 0.001% in gnomAD^8^ and < 0.01% in DDD, inherited, heterozygous variant that was either 1) a PTV or missense variant in a dominant DDG2P gene with a LoF mechanism, 2) a PTV or missense variant in a recessive DDG2P gene with a LoF mechanism or 3) a PTV or missense variant (MPC ≥ 2) in a constrained gene (pLI > 0.9). We used the DDG2P list downloaded on August 20, 2020, and focused on genes that were confirmed/probable DD genes.

In the first analysis (Figure 2), we compared genetically-predicted expression between probands with a putatively damaging variant in one of the aforementioned categories with their transmitting parent. Specifically, we tested (using a one-sided paired *t*-test) whether undiagnosed NDD cases carrying a variant in a given class had lower predicted gene expression than their parent who transmitted the variant. We only compared gene expression for one proband with one parent for one gene with a putatively damaging variant. If a proband inherited more than one putatively damaging variant, either from the same or both parents, a unique proband-parent-gene combination was selected at random with an equal probability of selection.

In the second analysis (Figure 3; Table 1), we calculated the percentile ranks of genetically-predicted gene expression values, per gene, in both cortex and whole blood, across undiagnosed NDD probands with putatively damaging variants in the gene and UKHLS controls. For each gene, we extracted the rank of genetically-predicted expression for cases carrying a variant in a given class, as well as the controls’ ranks. We then aggregated the ranks across genes and conducted a one-sided Wilcoxon rank test to test whether these ranks were lower in the variant-carrying cases compared to controls.

### Identifying undiagnosed probands with outlier expression in DDG2P genes

For each DDG2P gene, we identified undiagnosed NDD probands that had predicted gene expression at least three standard deviations above or below the mean predicted gene expression in controls from brain cortex or whole blood. We then conducted a Fisher’s exact test to test whether the number of cases with extreme levels of predicted gene expression was significantly different from that in controls.

## Data Availability

The DDD data are available in the European Genome-Phenome Archive (EGA). These include the exome sequence data (EGAD00001004389), phenotypic and family descriptions (EGAD00001004388), CoreExome array data (EGAD00010001598, EGAD00010001600, EGAD00010001604) and Global Screening Array data (EGA upload in progress, dataset numbers to be confirmed). The UKHLS genotype data are also available on EGA (EGAS00001001232).

## Acknowledgements

We thank the families and their clinicians for their participation and engagement, and our colleagues at the Wellcome Sanger Institute who assisted in the generation and processing of data, including the Human Genetics Informatics core.

The DDD study presents independent research commissioned by the Health Innovation Challenge Fund (grant number HICF-1009-003), a parallel funding partnership between Wellcome and the Department of Health, and the Wellcome Sanger Institute (grant number WT098051). This study makes use of DECIPHER^41^, which is funded by the Wellcome Trust. The full acknowledgements can be found at www.ddduk.org/access.html. Sanger investigators are currently funded by Wellcome grant 220540/Z/20/A.

We used data from UKHLS, which is led by the Institute for Social and Economic Research at the University of Essex and funded by the Economic and Social Research Council (grant number ES/M008592/1). The data were collected by NatCen.

## Additional Information

H.V.F. is an author of Oxford Desk Reference: Clinical Genetics & Genomics. M.E.H. is a consultant for AstraZeneca, and is a non-executive director of, consultant to, and holds shares in Congenica.

## Author contributions

Conceptualization, E.M.W., K.E.S., and H.C.M.; Formal Analysis, E.M.W.; Investigation, E.M.W., V.K.C., K.E.S., H.C.M., M.E.H.; Resources, C.F.W., H.V.F., M.E.H.; Data Curation, R.Y.E.; Writing - Original Draft, E.M.W. and H.C.M.; Visualization, E.M.W.; Supervision, H.C.M. and M.E.H.

## Notes

### Author Declarations

The DDD study has UK Research Ethics Committee approval (10/H0305/83, granted by the Cambridge South REC, and GEN/284/12 granted by the Republic of Ireland REC.

