## Supplementary Information for "Investigating the role of common *cis*-regulatory variants in modifying penetrance of putatively damaging, inherited variants in severe neurodevelopmental disorders"

#### Supplementary Methods

##### Principal components analysis on DDD individuals genotyped on the Global Screening Array

We identified individuals of European ancestries by projecting the DDD GSA samples onto 1,000 Genomes phase 3 individuals[20] (N = 2,548) using the smartpca function from EIGENSOFT version 7.2.1[64,65] (Supplementary Figure 5). We first filtered SNPs for MAF > 5% and Hardy-Weinberg Equilibrium p-value > 1 x $10^{-6}$. We LD-pruned SNPs pairwise $r^2$ > 0.2 in batches of 50 SNPs with sliding windows of 5 and removed 24 regions with high or long-range LD, including the HLA[66]. We then identified a subset of DDD samples who projected onto European-ancestry samples from 1,000 Genomes (PC2 > 0.0175), leaving 9,534 loosely-defined European-ancestry DDD individuals. We then ran another PCA with unrelated individuals within the loosely-defined European-ancestry subset, projecting related individuals once again using smartpca[64,65]. We used UMAP on the first 10 PCs of this PCA to further subset the loosely-defined European-ancestry individuals to a homogeneous ancestry group (presumed to be of GBR ancestries) and plotted PCs to be sure there was no remaining heterogeneity.

##### Principal components analysis on DDD individuals genotyped on the Global Screening Array or Omni chip combined

We identified trios in which all three individuals had GBR ancestries (as determined in the PCA of individuals genotyped on the GSA chip described above), or identified in Niemi *et al.*[15] as having GBR ancestries (from a PCA of individuals genotyped on the OmniExpress Chip). This left 11,227 individuals. Of these, we selected 3,835 unrelated probands to conduct a PCA, and projected remaining family members, using smartpca[64,65] (Supplementary Figure 6).

### Supplementary Figures

#### Supplementary Figure 1

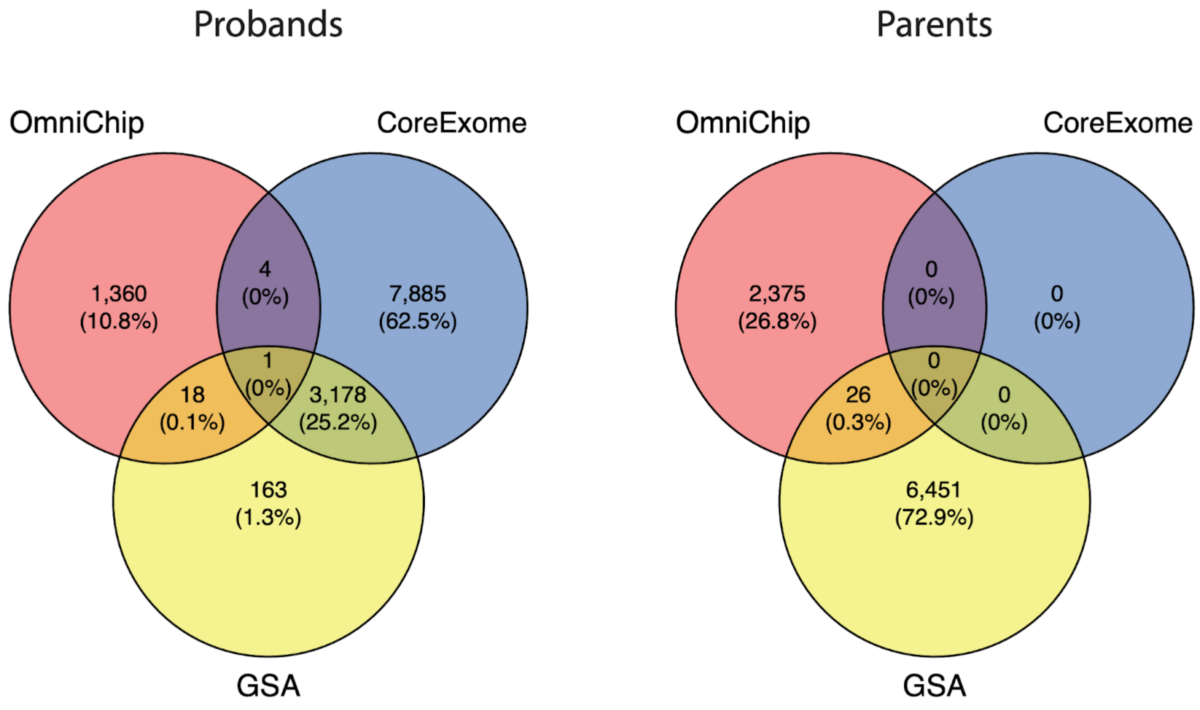


Venn diagram showing which DDD individuals were genotyped on which arrays.

Supplementary Figure 2

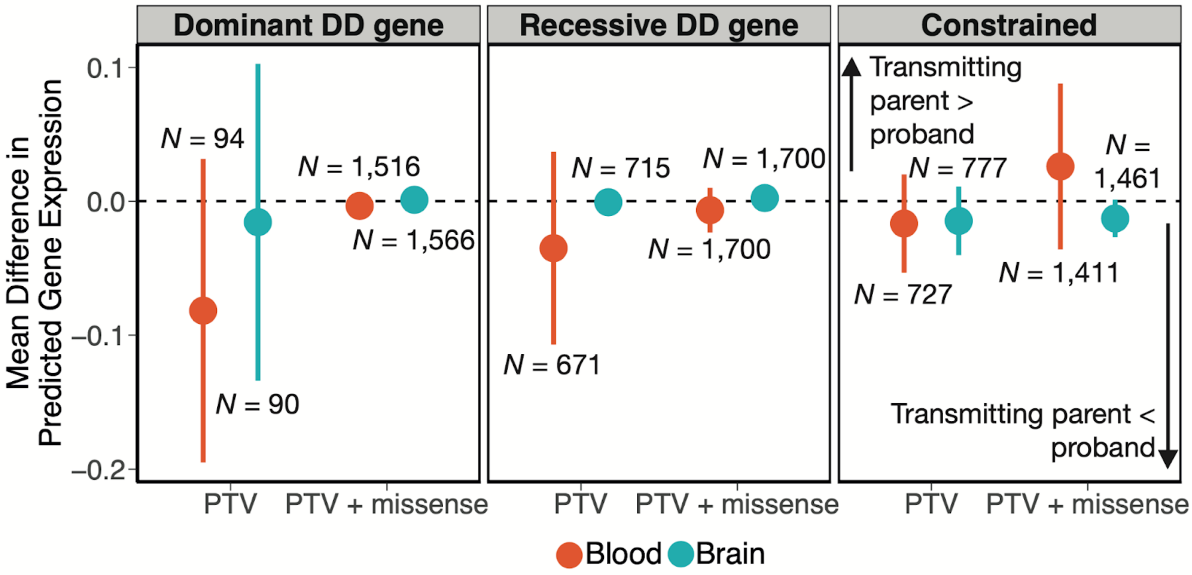


Mean difference (parent - child) in predicted gene expression between parents transmitting putatively damaging variants (with MAF < 0.1%) and their children with an undiagnosed NDD, with 95% confidence intervals. This is the same analysis as Figure 2B except with a less stringent MAF cutoff. *N* is the number of unique child-parent pairs. The difference in predicted gene expression is in the inverse quantile-normalised number of reads per kilobase of transcript per million mapped reads (RPKM). The three panels show results for putatively damaging variants in three different sets of genes: dominant DD-associated genes with a LoF mechanism (left), recessive DD-associated genes with a LoF mechanism (middle) or constrained genes (pLI > 0.9) (right). Red and blue dots represent results from genetically-predicted gene expression imputed from whole blood and cortex, respectively. We show estimates considering only PTVs, as well as PTVs and missense variants (with MPC ≥ 2) together.

Supplementary Figure 3

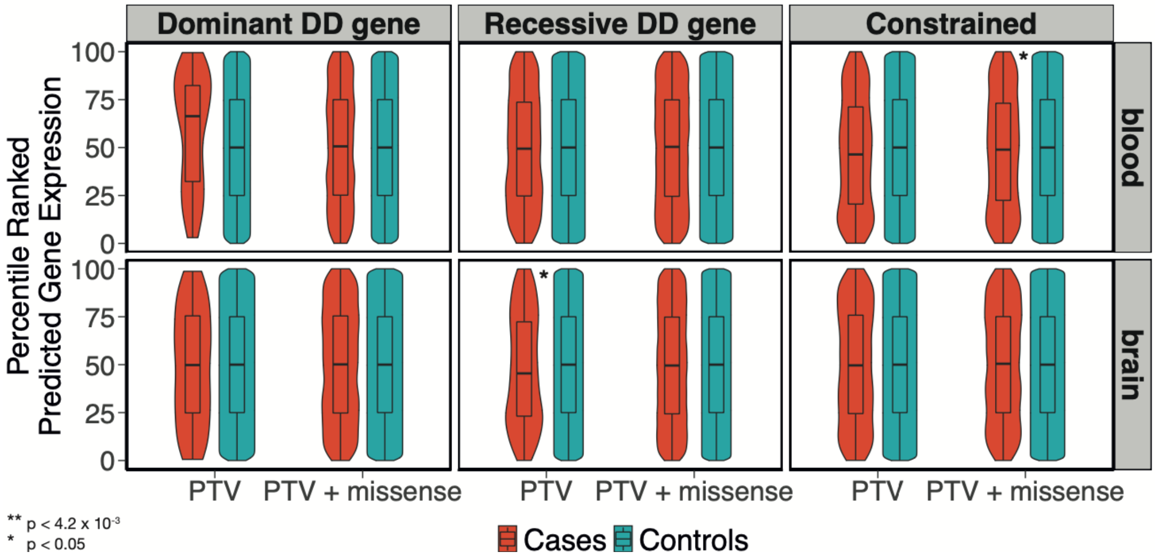


Violin and box plots of percentile-ranked genetically-predicted expression of genes harboring putatively damaging variants (MAF < 0.1%) in undiagnosed NDD cases, compared to controls. This is the same analysis as Figure 3, but with a less stringent MAF cutoff. Vertical lines of the box plot indicate the range and horizontal lines indicate the lower quartile, median and upper quartile. The p-value is from a one-sided Wilcoxon test assessing whether cases are lower than controls.

Supplementary Figure 4

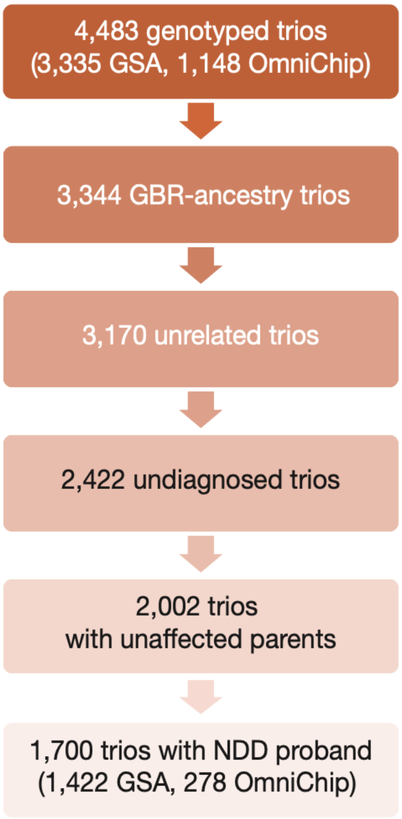


Flow diagram of DDD trio filtering.

Supplementary Figure 5

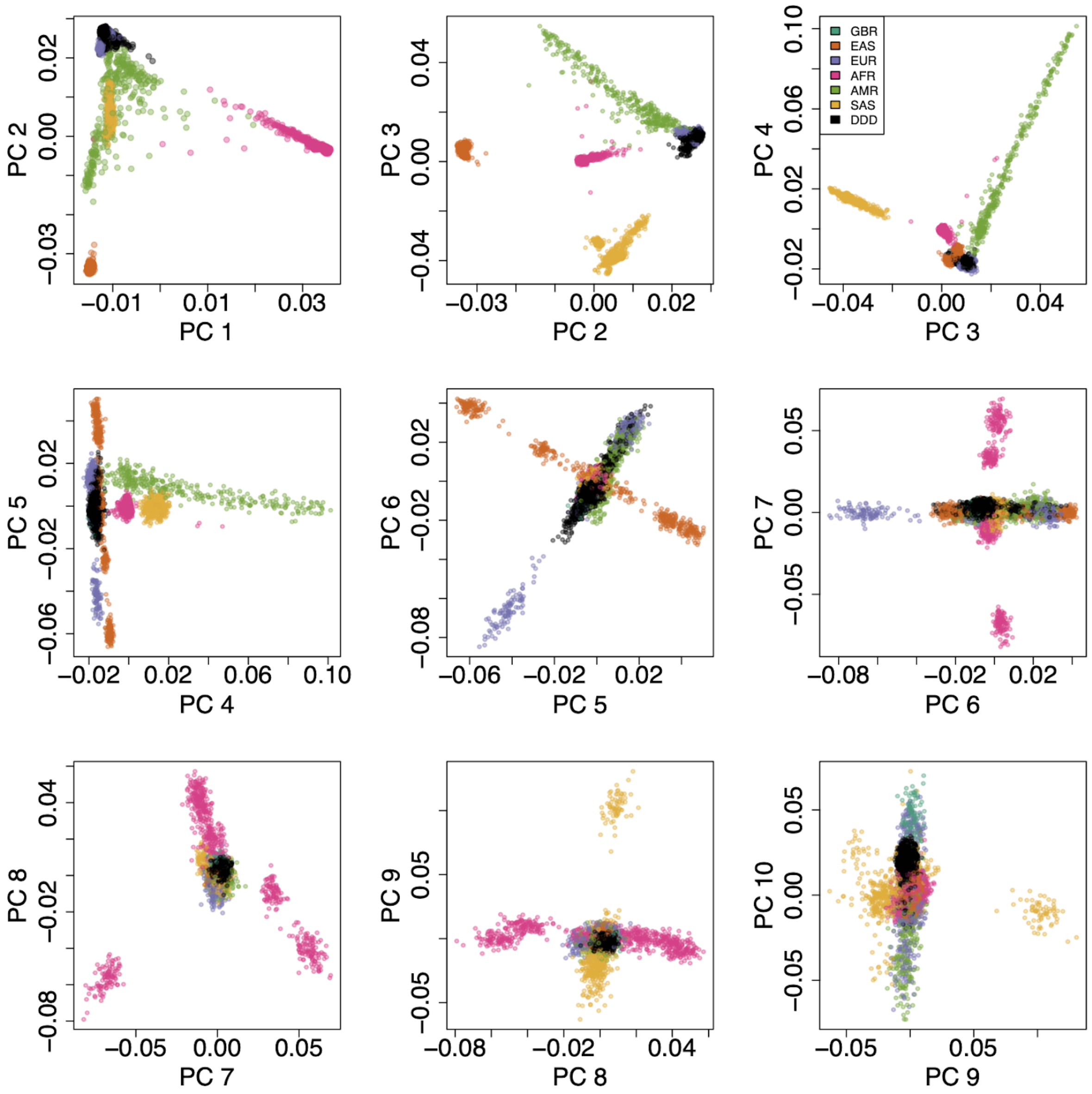


Principal components of genotypes of DDD individuals on the GSA array (N = 9,572) and 1,000 Genomes phase 3 (N=2,548). DDD participants are in black, and 1,000 Genomes participants are coloured by superpopulation, with the exception of GBR individuals, which form a subpopulation of the EUR superpopulation. GBR: Great British, EAS: East Asian, EUR: European, AFR: African, AMR: Admixed American, SAS: South Asian.

Supplementary Figure 6

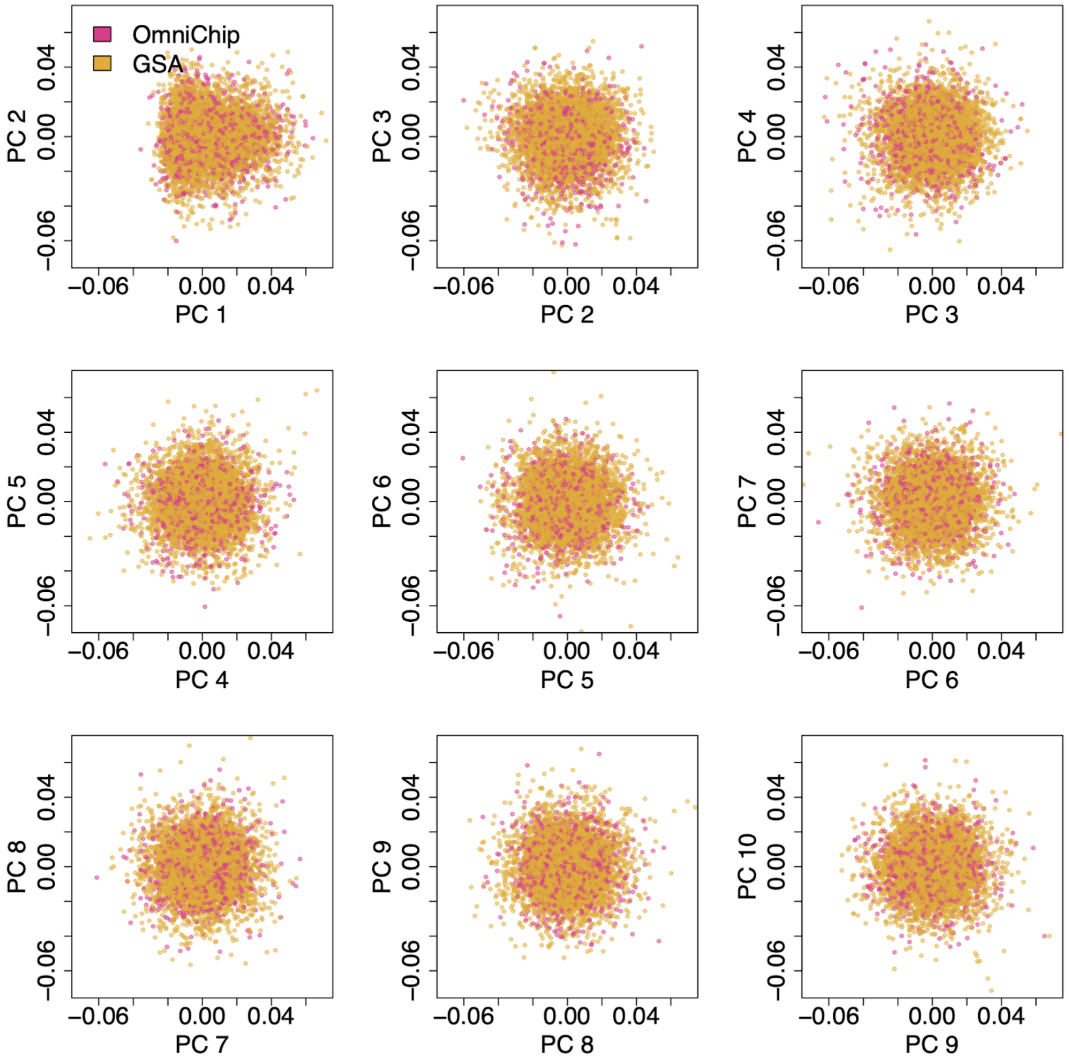


PCA of GSA chip individuals inferred to have GBR ancestries and OmniExpress chip GBR individuals from Niemi *et al.*[15].

### Supplementary Tables and Data

#### Supplementary Table 1

Number of individuals and SNPs remaining from a pilot batch of DDD individuals (N=1,152) genotyped on the GSA array and a larger second batch of individuals genotyped on the GSA array (N=8,698) after various quality control filters.

#### Supplementary Table 2

Number of individuals and SNPs remaining after merging a pilot batch of DDD individuals (N=1,152) genotyped on the GSA array and a larger second batch of individuals genotyped on the GSA array (N=8,698) after various quality control filters.

#### Supplementary Table 3

Positive predictive values for different classes of variants reported into DECIPHER[41] from DDD probands. These were used to help decide which probands were potentially diagnosed versus undiagnosed (see Methods).

#### Supplementary Table 4

Count of individuals and variants after various filtering steps to conduct PCA within European-ancestry individuals genotyped on the GSA array, and count of individuals and variants after selection of the samples from a homogeneous European population (inferred to have GBR ancestries).

#### Supplementary Table 5

Summary of quality control metrics applied to whole-exome sequence data from DDD.

#### Supplementary Data 1

Summary statistics from TWAS in cortex. The columns are as follows:
**ensembl_gene_id**: gene ID from Ensembl human genome build 38
**gene**: gene symbol
**chr**: chromosome
**b38_start**: gene start position in human genome build 38
**b38_start**: gene end position in human genome build 38
**beta**: effect size from TWAS (regressing predicted gene expression on case/control status, controlling for 20 PCs)
**se**: standard error for beta
**z**: z statistic for beta
**p**: p-value for beta
**log_p**: $-\log_{10}$ p-value for beta
**p.adj.fdr**: FDR-adjusted p-value for beta

#### Supplementary Data 2

Summary statistics from TWAS in blood.The columns are the same as for Supplementary Data 1.
